# Diabetes Technology Experiences of Young People Living with Type 1 Diabetes Mellitus and their Parents: Hybrid Theoretical Foundation Guided Analysis

**DOI:** 10.1101/2021.11.08.21265793

**Authors:** Nicola Brew-Sam, Madhur Chhabra, Anne Parkinson, Adam Henschke, Ellen Brown, Lachlan Pedley, Elizabeth Pedley, Kristal Hannon, Karen Brown, Kristine Wright, Christine Phillips, Antonio Tricoli, Christopher J. Nolan, Hanna Suominen, Jane Desborough

## Abstract

An important strategy to understand young people’s needs regarding technologies for Type 1 Diabetes Mellitus (T1DM) management is to examine their day-to-day experiences with these technologies. This study aimed to describe T1DM youth and their caregivers’ experiences and preferences regarding insulin pumps, sensor technologies, and related communication technologies based on a hybrid theoretical foundation, as well as to describe derived ideal device characteristics. Sixteen face-to-face interviews were conducted with young people and their parents. Data analysis included data-driven thematic analysis followed by theory-driven analysis (Unified Theory of Acceptance and Use of Technology; value sensitive design). Initial themes derived from the interviews included aspects of self-management, device use, technological characteristics, and feelings associated with device types. Interview findings were congruent with factors from the two theories. Discussions around ideal devices focused on reliability, flexibility, and automated closed loop systems that enabled an independent life for young people and alleviated parental anxiety. Reality deviated from expectations, with inaccuracy problems and technical failures reported. Technologies for diabetes self-management require continual advancement to meet the needs of young people with T1DM and their caregivers. Understanding experiences and challenges with devices enabled us to identify theory-supported device characteristics useful for the designing of improved technologies.

**Extended abstract (structured):** *Background:* An important strategy to understand young people’s needs and preferences regarding technologies for Type 1 Diabetes Mellitus (T1DM) management is to examine their day-to-day experiences with these technologies.

*Objective:* This study aimed to describe T1DM youth and their caregivers’ experiences and preferences regarding insulin pumps, sensor technologies, and related communication technologies based on a hybrid theoretical foundation, as well as to describe derived ideal device characteristics.

*Materials and Methods:* Sixteen face-to-face interviews were conducted with young people with T1DM and their parents about their diabetes technology use. A combination of data-driven thematic analysis in a first stage, and theory-driven analysis in a second stage was used to incorporate in-depth study analysis and existing theoretical literature. Relevant literature included technology adoption (Unified Theory of Acceptance and Use of Technology/UTAUT) and value sensitive design (VSD) models. Based on this approach ideal device characteristics for young people with T1DM were summarized.

*Results:* Initial themes derived from the interviews included aspects of diabetes self-management, device use, and specific device-related technological characteristics and feelings associated with the specific device types (continuous glucose monitoring, insulin pump, flash glucose monitoring). The interview data delivered information congruent with all UTAUT and VSD factors except for one (privacy). Discussions around ideal diabetes devices focused on reliability, flexibility, and automated closed loop systems that enabled an independent and normal life for adolescents, and alleviated parental anxiety. However, in line with the previous systematic review, the interview analysis showed that reality deviated from these expectations, with inaccuracy problems reported for continuous glucose monitoring devices, and technical failures occurring in both continuous glucose monitoring devices and insulin pumps.

*Conclusions:* UTAUT and VSD approaches were found useful as a combined foundation for structuring our study findings. Technologies for diabetes self-management require continual advancement to meet the needs and expectations of young people with T1DM and their caregivers. Understanding their experiences, as well as challenges with the devices, enabled us to identify theory-supported ideal device characteristics that can be useful in the designing and developing of improved technologies.

## Introduction

Type 1 Diabetes Mellitus (T1DM) is an autoimmune condition diagnosed in children and adolescents often at an early age [1]. It requires life-long self-management, including blood glucose monitoring, adherence to insulin regimens, adjustments to lifestyle, including diet and exercise, and for many, psychological management [2]. Advanced diabetes technologies such as insulin pumps, continuous glucose monitoring systems (CGMs), or closed loop systems, have been found to improve self-management and quality of life for young people with diabetes [3]. Moreover, the use of such technologies can help reduce the risk of acute and long-term diabetes complications [4].

An important strategy to understand young people’s needs and preferences regarding technologies for T1DM management is based on an examination of their day-to-day experiences with these technologies. The analysis of experiences with technologies also serves to identify specific technology usage perceptions, decisions and behaviors [5, 6]. Moreover, technology experience can enrichen co-design approaches using feedback received based on technology use [7]. The importance of understanding experiences with diabetes technologies in young people with T1DM has led to a variety of studies conducting research on this topic [8]. The majority of these studies chose exploratory research designs to investigate experiences with diabetes technologies in young target groups [8]. To follow up and to expand this existing body of research, available theoretical knowledge from models and theoretical approaches can be combined with exploratory research approaches. Previous studies have shown that a stronger focus on theory can strengthen and advance research outputs [9, 10]. Incorporating theoretical knowledge into the study of experiences and preferences of young people regarding diabetes technologies can promote a well-grounded foundation for future research in this area, and can inform the development of improved diabetes technologies for young people on a sound basis.

> *We aimed to describe the experiences and preferences of young people with T1DM and their caregivers regarding insulin pump, sensor, and communication technologies based on a hybrid theoretical foundation, as well as to summarize derived ideal device characteristics*.

## Materials and Methods

We conducted 16 interviews with young people with T1DM and their parents about their diabetes technology use. The interviews were conducted face-to-face, either offline or online, by four researchers (NBS, MC, AP, JD) between December, 2019 and July, 2020 (12 offline face-to-face interviews, four online video calls due to the onset of COVID-19) until data saturation was reached. Interviews were between 20 to 30 minutes duration. The interview protocol was informed by existing literature and developed in collaboration with young people living with T1DM who were members of our research team. It contained five general questions about managing diabetes with technological devices, device types used, decision making, and encountered challenges, and was extended to include important aspects repeatedly mentioned in the interview process. All interviews were professionally transcribed. Ethical approval was obtained by the Australian National University’s Human Ethics Committee and ACT Health Human Research Ethics Committee (HREC, Australia) in October 2019 (2019.ETH.00143; 2019/ETH121700). In addition, a variation for online interviews was approved by the same committees in April, 2020. All interviews were based on informed consent.

For data analysis, we used a combination of data-driven thematic analysis in the first stage, and theory-driven analysis in the second stage in order to pay respect to both rich data from the interviews and the existing literature. The first stage comprised a qualitative data-driven thematic analysis approach based on Braun and Clarke [11] which was used to extract the main themes discussed in the 16 interviews. Two researchers (NBS, MC) conducted the coding for an initial codebook, and ongoing discussions between four researchers (NBS, MC, AP, JD) were held throughout the data analysis process to ensure result validity. In the second stage, the alignment of the resulting themes was analysed in relation to the key factors from the Unified Theory of Acceptance and Use of Technology (UTAUT) [12] and value sensitive design (VSD) factors [13] in a theory-driven analysis. Both theoretical approaches – technology adoption and technology design – are highly relevant as a foundation for the examination of experiences with diabetes technologies. Thus, an integration of knowledge from both offered a step towards a comprehensive understanding of user experiences with diabetes technologies.

### Hybrid theoretical foundation of theory-driven analysis

The application of technology adoption models has been recognized as a sound foundation for the examination of health technology usage decisions and behaviors. The UTAUT is a model incorporating elements from eight previous well-established models [12], and has been applied in recent studies examining acceptance of information and communication technologies (ICT) by diabetes patients [14, 15] and/or by healthcare professionals [16]. The UTAUT illustrates that the key *factor experiences with technologies* (**Table 1**) moderates associations between various antecedent factors and usage intention [12] or technology use [17], and influences technology uptake [18, 19]. Technology adoption models are subject to certain limitations due to their binary logic of technology adoption [20] and assumption of rational behavior [21]; these are at odds with the principles of a complex self-managing ecosystem in which users with varying needs, desires, and interests make decisions and act within a sociocultural context [13]. VSD approaches offer an additional holistic approach that integrates users’ values and life circumstances into an examination of interaction with technologies [22]. VSD is used to conceptualize and identify users’ values and to design technologies in accordance with these values [13]. Dadgar and Joshi [13] summarize 12 values and four system features relevant for the design of diabetes technologies from their study of adults with T1DM (**Table 1)**. They acknowledge that patients’ technology use is embedded in self-management activities and in relationships with family, friends, and healthcare providers.

**Table 1.** Explanation of UTAUT and VSD Factors

| Model | Factor | Explanation |
| --- | --- | --- |
| UTAUT core determinants | Effort expectancy | The ease of the technology use |
|  | Facilitating conditions | Include infrastructure (e.g., organizational or technical) supporting the technology use |
|  | Performance expectancy | The extent to which the user believes that using the technology will improve his/her performance |
|  | Social influence | The degree to which someone thinks that important others believe he/she should (not) use the technology |
| UTAUT moderators | Gender/age | – |
|  | Usage experience | The previous experience with the technology |
|  | Voluntariness of use | The degree to which the technology use is perceived as carried out of free will |
| VSD system features | Connectivity | Features that allow the user to interact and share information with others |
|  | Data analysis | Features that allow the user to make sense of data over time |
|  | Data retrieval and storage | Features that allow the user to access, store, log, and track data |
| VSD values | Accessibility | The system's availability and portability |
|  | Accountability and autonomy | Self-responsibility for habits and care performance, with independent behavior and decision making |
|  | Compliance | Adherence, following diabetes care plan |
|  | Dignity | Sense of pride and self-respect (impacted by negative outcomes or unfair treatment for performance) |
|  | Empathy | Desire to be understood by others |
|  | Feedback | Responses by others or technology |
|  | Hope and joy | Motivation to achieve future-oriented expectations and personally valued goals, also including joy in life |
|  | Privacy | Information protection when sharing sensitive (health) data |
|  | Sense-making | Ability to give meaning to data |
|  | Trust | Expectations of trust in technology, oneself, others |

*Notes.* UTAUT factor explanations based on Venkatesh, Morris [12], VSD factor explanations based on Dadgar and Joshi [13]

Based on the inclusion of both theoretical approaches in our data-analysis, conclusions about experiences with diabetes technologies and preferences in young people with T1DM were drawn and discussed in the context of previous literature. Ideal device characteristics for young people with T1DM were then summarized.

## Results

### *Study Sample and Devices* Used

The sample included 16 young people with T1DM (female n=7, male n=9) between 12 and 17 years of age^1^, and accompanied by a parent (mother n=11, father n=4, both=1). The young people had been diagnosed with T1DM from one to 14 years prior to this study. Thirteen participants used an insulin pump (Tandem t:slim n=3, Medtronic n=10), 14 used a CGM (Dexcom n=11, Medtronic Guardian n=3), and one used a flash glucose monitor (FGM, Freestyle Libre). All participants either used an insulin pump (n=2), a CGM (n=3), or a combination of both (n=11). The flash glucose monitoring system was used in addition to the pump. Additional devices used included an Apple watch (previously used n=2, current use n=1, planned n=1), diabetes apps on smartphone/smart devices (various), and glucose and ketone meters (n=16). The study sample overview can be found in **Table 2**.

**Table 2.** Study Sample Overview

| IP | Mode | Gender | Year of diagnose (range) | CGM | Insulin pump | Other technology |
| --- | --- | --- | --- | --- | --- | --- |
| 1 | Face to face | M | 2016-2020 | Dexcom G5 | No pump | – |
| 2 | Face to face | F | 2011-2015 | Dexcom | Tandem T: Slim T3 | Previously apple watch |
| 3 | Face to face | M | 2011-2015 | No CGM/<br>previously<br>Dexcom G5 | Medtronic | – |
| 4 | Face to face | M | 2016-2020 | Dexcom | Tandem T: Slim | Free-style Libre,<br>thinking about Apple<br>watch |
| 5 | Face to face | M | 2006-2010 | Dexcom | Medtronic 640G | – |
| 6 | Face to face | M | 2006-2010 | Dexcom | Medtronic | Apple watch |
| 7 | Face to face | F | 2006-2010 | Dexcom G5 | Medtronic | – |
| 8 | Face to face | F | 2016-2020 | No CGM | Medtronic | – |
| 9 | Face to face | M | 2006-2010 | Medtronic/<br>Guardian | Medtronic | – |
| 10 | Face to face | F | 2006-2010 | Dexcom | Medtronic | – |
| 11 | Face to face | M | 2011-2015 | Dexcom | Medtronic | – |
| 12 | Face to face | F | 2016-2020 | Dexcom | Tandem T: Slim | – |
| 13 | ZOOM | M | 2011-2015 | Medtronic/<br>Guardian | Medtronic 640G | previously Dexcom and<br>Apple watch |
| 14 | ZOOM | F | 2016-2020 | Dexcom G5 | No pump yet, t: slim<br>X2 planned | – |
| 15 | ZOOM | M | 2006-2010 | Medtronic/<br>Guardian | Medtronic 670G | – |
| 16 | ZOOM | F | 2016-2020 | Dexcom G5 | No pump | – |

*Note.* Participants between 12 to 17 years of age; each young participant was accompanied by a parent/the parents

### Interview Themes and Alignment with Theoretical Factors

Initial themes identified from the interview data included information related to (1) sociodemographic characteristics, (2) medical diabetes and diabetes self-management (e.g., diagnosis, family members with diabetes, diabetes education, hypo/hyperglycemic awareness and events, style of self-management), (3) device use (e.g., types of devices used, length and frequency of use, preferences), and (4) specific device type related technological characteristics and feelings associated with the specific device type used (CGM, insulin pump, FGM) (**Appendix 1**).

In the following, the findings focus on the alignment of the initial themes with UTAUT and VSD factors, and are reported in accordance with these factors (alignment and data excerpts in **Appendix 2**). Participants’ statements are cited with interview participant numbers (e.g., young interview participant 1 displayed as IP1), and their parents’ statements are indicated with an additional “P” following the number (e.g., the parent of the young interview participant 1 displayed as IP1P).

### UTAUT Factors

#### Performance expectancy

Participants described how CGM and pump technologies contributed to success in T1DM self-management, and how the use of the devices made self-management easier. CGM improved and facilitated blood glucose tracking (IP14; IP16), allowed the young people to have breaks from diabetes management (IP13P), and led to *“satisfaction of seeing it successful”* (IP1). The direct connection of CGMs to phones was considered a major benefit (IP7). The insulin pump assisted in stabilizing blood glucose levels (IP14; IP15), reduced the use of needles (IP2, IP6,) and allowed flexible eating (IP2; IP6; IP15), and was viewed as convenient, enabling a normal life (IP5P). Reduction in the use of injections by using a pump was perceived as a major advantage for small children (IP5P). Avoiding calculations was considered another benefit of the insulin pump (IP12P). Most of the young people expressed appreciation of having the devices, and this was also highlighted by one parent (IP6P).

#### Effort expectancy

The CGM was perceived as easy to use and to put on (IP16). It did not require constant *“fiddling”* (IP13) and saved time (IP1; IP5). The phone could quickly be checked instead of using finger pricks (IP2; IP6; IP7; IP11; IP12). Flash glucose monitoring (FGM) was perceived as easy to use because *“you just swipe it and you get the number”* (IP4), as was the insulin pump (IP8P). Participants mentioned that they had to *“calculate the carbs”* for calculating the insulin dose delivered through the pump (IP3; IP4P; IP8P). “*When you just click that, so you can bolus there, put a basal on, like all that cool stuff”* (IP7). The pump was considered easier to handle than insulin pens (IP5P; IP6; IP9), also due to data storage options (IP6) and *“pre-programmed”* calculations (IP3).

> *“I think the biggest thing when [name] got the pump for us as a family, it has made it much easier because*…, *on a long car journey, we’ve got two older children, so they’d be constantly ‘can we have some foodã’ And I’d be like ‘no, because [name] has got to have another injection’. So once he got the pump, ‘yeah, sure you can have some. Just dial up some more insulin, [name]*.*’” (IP6P)*

Difficulties charging devices in some situations were mentioned (IP6P) which led to the use of insulin pens in certain circumstances (IP6P). However, in most situations the pump could be used (see *accessibility*).

#### Social influence and voluntariness of use

Physicians recommended CGMs (IP6; IP14) and insulin pumps (IP1P; IP2P; IP16P). Young people with T1DM paid particular attention to the devices used by their peers with T1DM (IP14). This helped them make decisions regarding their own devices (IP16).

> *“I ask them about their pumps, because they’ve all got the same pump, so I talk to them about it and try and get, like, what they think about it*.*” (IP14)*

Social influence on device decisions was closely related to voluntariness of use, reflecting the degree to which the technology use was perceived to be volitional [12]. Device decisions were taken by parents for very young children (IP10P), while adolescents reported they made their own informed decisions, or together with their parents (IP3P). The parents of all participants were closely involved in diabetes management as they were all minors. Nevertheless, some young people decided against insulin pumps (IP1; IP16) even though their physicians had recommended it, and parents accepted these decisions (IP16P; also see accountability).

#### Usage experience

Participants reported that diabetes diagnosis was stressful, especially the early days after diagnosis, and that they had tried to learn about the disease and about how to operate the devices (IP3-5P). After that, a self-management routine was established and self-management became easier, especially for participants who had used the devices for longer or had been living with diabetes for an extended period of time (IP3P; IP6P).

> *“She was only a baby*, … *with rotavirus*… *that was the trigger. Took her to the doctors, he said ‘oh look* … *it could be diabetes’* … *I had no idea what he was talking about He phoned me that night and he said ‘we need you down the hospital straight away’. And at that point, my life changed*.*” (IP10P)*

Participants also reported trialling multiple devices until the best self-management solution was found to keep blood glucose in range (IP13; IP15).

> *“Before that he tried the Dexcom CGM, and before that he tried the Medtronic Guardian one*… *it didn’t really work, so that’s why we went to the Dexcom, and then since he’s been on this new pump, then we went back to the Medtronic one*.*” (IP15P)*

#### Facilitating conditions

Health insurance and subsidizing schemes were reported to impact device use and choice, and there were waiting periods for chronic conditions and device replacements (IP2P; IP3P; IP7P; IP8P; IP13-16P), and delays in technology release processes (IP5P; IP12P; IP14P). The release of new pump features was welcome, but at the same time parents expressed concerns about effects of new features on self-management, such as those overriding basal adjustment (IP10P). There was a desire for improved funding options to pay for devices (IP5P). The high cost of devices – especially the insulin pump – was criticized by several participants (IP2P, IP4, IP5P, IP7), which also led to fear that devices could break (IP5P), or attempts to extend a device’s lifespan (IP2P).

*“We had to wait until our health insurance covered it. Because they’re expensive. And so now we’re waiting to get the Medtronic sensor, because it’s covered with one of the rebates or whatever that the government do for people under 21, but it’s not covered for my older [child]*… *It’s very expensive, it’s thousands of dollars a year*.*” (IP7P)*

Two participants mentioned problems with customer service, provided through a company hotline, for both CGM devices (IP2P) and for insulin pumps (IP13P). In contrast to some difficulties with device customer service, most participants mentioned good hospital infrastructure with ongoing support provided by the diabetes healthcare team including training about how to use an insulin pump (IP14).

> *“That’s the staff at the Canberra Hospital, they’re brilliant*… *There’s the paediatric diabetes team at the Canberra Hospital … the diabetic educators. They’re great*.*” (IP13P)*

A hospital hotline was available for the families connecting them to diabetes educators or on-call registrars (IP4; IP14P), as well as email contact to doctors, educators, and dietitians (IP14; IP14-16P). Mobile numbers for the endocrinologist were also provided (IP14P), and phone consultations in addition to face-to-face consultations were possible during the COVID-19 pandemic (IP14P). Parents valued face-to-face support from the healthcare team (IP15-16P). For some participants who had been treated in both rural and urban clinics, the rural clinics were reported to deliver less efficient consultations than urban clinics (IP2P; IP15P), and communication between the healthcare team in a rural setting was criticized by one participant (IP15P). The interview data pointed to the possibility that quality of diabetes support might also vary between general practitioners/paediatricians and diabetes specialists (IP13P; IP15P).

In addition to healthcare support, Facebook groups for parents of children with T1DM (or other social media) were used for non-medical queries (IP13P). Support by school personnel such as teachers, was described as being important for safe diabetes management outside of home. Most of the young participants with T1DM reported teachers were supportive regarding device use and diabetes management in class (IP5; IP6P; IP7; IP8-9P; IP13P, IP13-15; IP16P), however some highlighted problems with relief teachers (IP2; IP4), or inappropriate behaviors by uninformed teachers or the school (IP3).

*“There was one time where a teacher asked to take the pump*… *well, one of my diabetic friends that goes to that school, he got his pump off him for the day, which probably wasn’t good*.*” (IP4*)

School management plans regarding device use in school were agreed on together with the healthcare team (IP3P; IP14), and in close cooperation with parents (IP6P). CGMs were particularly perceived as helpful in the school environment (IP16P).

### VSD Factors – System Features

#### Connectivity

While the pump could not directly be connected to a phone for easier operation (IP14), and direct data sharing from the pump was not possible (IP2), most participants used a CGM to make *the “devices talk to each other”* (IP14P) (reported also by IP2; IP3P; IP4; IP12). They tried to achieve a closed loop system, with partial success (IP7P), connecting the phone, the CGM, and the pump (IP2), some used an Apple watch in addition (previous use: IP13/IP13P; current use: IP6; IP2). While the Medtronic system (CGM plus pump) had the *“suspend when low”* function (IP2P; IP5P), the Dexcom/pump combination allowed data to be shared with several other devices (IP7) but did not provide the suspend function and vice versa (IP1; IP5P). A combination of both was wished for (IP1P; IP5P; IP9P). It was perceived as difficult to choose one pump or CGM system, *as “they all have their pros and cons. A bit like Ford and Holden [cars]”* (IP2P). Depending on the chosen system, patient data could be automatically accessed by the healthcare team for some patients, for example through the Dexcom Clarity (IP4; IP13P; IP14; IP16P), others had to upload their data to a cloud system to share them with the healthcare team (e.g., Medtronic CareLink, IP15P). Apart from the healthcare team, Dexcom data were mostly shared within the family (IP1P; IP4-5P; IP7; IP12P; IP13-14), particularly for younger children (IP15P). Parents especially valued the sharing option (including alarms) as a *“safety net”* (IP1P; IP7P; IP9P; IP16P). This option also made the Dexcom the most popular CGM in the study sample (IP1).

#### Data analysis, data retrieval and storage

Participants reported that their endocrinologists used the transmitted CGM data to calculate an average value resembling the HbA1c, especially when HbA1c testing was not possible (IP16P). However, this was described as *“not always a hundred percent accurate”* (IP14P). Moreover, participants valued weekly summaries (IP4; IP7P; IP14), data trends (IP1P; IP14), and other graphical output/visualisation options of the CGM (IP5P).

To access or retrieve diabetes information, parents used diabetes information websites (IP13P), Facebook groups for people with T1DM (IP1P; IP13P), and Google (IP4P). For access to blood glucose data or information about food, young people used glucose tracking apps and food database apps (IP4).

> *“So when I’m on my phone I’ll quickly switch to that [app] and check it. So then when I turn on the phone, I just glance at it and do my business and that before I turn it off, I just check it again*.*” (IP1)*

Data storage was reported as a feature of insulin pumps and blood glucose meters (IP6).

### VSD Factors – Values

#### Accessibility

Accessibility was mentioned when young people and their parents described situations in which the technology required flexibility. This included diabetes management at night, during sports, in school, or when participating in sleepovers or camps. Devices facilitated attendance at camp or sleepovers (IP1P; IP15P), with the CGM and its data sharing options being more useful than the pump (IP6P; IP9). Some young people kept their CGMs on their bodies during sports and swimming (IP7; IP12; IP16), some took it off for swimming only (IP6). Water resistance of the pump was mentioned by one participant (IP4). The device tapes came off at times (IP1; IP2P; IP5; IP7-8; IP13-14), and better adhesives were requested (IP1; IP3; IP14P), as well as reduced device sizes to facilitate physical activity (CGM: IP1/IP3; pump: IP6-7 *“bulky”*). Participants expressed a desire for devices that were small but still effective (IP4), and for fewer devices that had to be carried (IP1P; IP5P).

> *“What is needed is an all in one device (CGM, insulin pump and control system) that doesn’t require tubes and can be controlled via an app with an algorithm that constantly regulates blood sugars that can operate as a closed loop system*.*” (IP14P)*

Taking the pump off during swimming (IP11; IP13) or sports (IP10; IP15) initiated a “panic mode” that *“you need to put it on silent otherwise your bag is making all sorts of wonderful noises”* (IP13P). Device alarms were reported to be challenging in various situations (IP9; IP15P), interfering with sleep (IP1-3P; IP12; IP13P) or activities in school. Alarms were perceived as embarrassing in school (IP1; IP3-4; IP15) which led to ignoring (IP2P; IP6P; IP9) or limiting them (IP7; IP15P), or turning them onto vibration/silent mode (IP2-4; IP15). Some parents tried to teach their children not to be ashamed of their devices (IP2P). Yet, at night, alarms created a feeling of safety (IP1; IP3-4P; IP14), especially for parents when the young people would sleep through them (IP1P; IP2; IP4; IP5P; IP6-7; IP8P; IP9; IP14P; IP16P). Alarms could be customized for different situations (IP1).

#### Accountability and autonomy

At night most parents reported taking over care of their children’s diabetes management (IP2P; IP5P; IP7; IP8P; IP10P; IP13P; IP16P) which related to perceptions of accountability and autonomy [13]. One participants stated that responsibility lay with the parents at night (IP1). Parents commonly transferred part of the responsibility to their children when they became teenagers (IP2P; IP15P), assisting where needed (IP2P; IP16P). At that stage, adolescents preferred some independence from their parents and freedom to make their own decisions (IP2-3P; IP3; IP5; IP9; IP10P), as they felt more confident and in control of their diabetes devices (IP14).

> *“I feel like you reach a point where we kind of know a bit more [than the doctor]*… *because we’re the ones experiencing it kind of every day*.*” (IP14)*

Some adolescents felt like role models for younger children with T1DM (IP4P). In contrast to young people, some parents had problems letting go of the responsibility, wishing to continue to share data (IP15P), which was at times perceived as intrusive by the young people (IP2) as they reported being fine without CGM data sharing (IP15). Yet, CGM data sharing also facilitated independence in some young people and reduced anxiety in parents when their control over the children was reduced (IP16P).

#### Trust

Independent management was associated with trust in the devices, affected by accuracy and device failures (IP8P). CGM technology was reported to be inaccurate at times (IP1-2; IP3P; IP8; IP14/IP14P; IP16P) for example when *“it*… *wears down”* (IP16P) (similar: IP11), with time lags occurring (IP1; IP3P; IP10-11; IP12-13P; IP16P), while the pump was mostly accurate and reliable (IP3; IP8; IP15). Technical device failures, such as blocked insulin tubing and other issues, were reported for both the pump (IP4; IP6; IP9; IP15) and the CGM (IP1-2; IP8P; IP10; IP13). Most participants used finger pricking as a backup option when they were uncertain about device accuracy, or to re-calibrate the device (IP1-6; IP8P; IP10; IP12; IP13P; IP14-15; IP16P). They also thought of other measures to improve the safety net such as a diabetes assistance dog (IP1P). Device calibration was perceived as difficult at times, for example taking paracetamol was experienced as affecting blood glucose readings and respective calibration (IP1-2; IP14). Several participants mentioned that they trusted their bodies and the blood glucose meter more than the CGM devices (IP1; IP5; IP8; IP14-15), telling them when hyper or hypo events occurred (IP3; IP10; IP12; IP13P). Trust in the healthcare team was equally relevant, as this gave the participants a feeling of safety in case they needed medical support. This was reported by almost all participants (see *facilitating conditions*).

#### Sense-making

Participants stated that their ability to make sense of the data and give meaning to it increased with advancing age, length of the disease, and independence in the young people with T1DM. Diabetes education and device training played a crucial role for understanding data and managing diabetes independently (IP14; IP16P). It was described as a gradual and individual process of learning how to best deal with the disease, its management, and device use (IP13). Graphical device outputs facilitated sense-making of numerical values (*see data analysis*). Management approaches were individual and solutions had to be adapted for each patient, with no one-size-fits-all solution available (IP14; IP15P). While some participants preferred multiple daily injections (MDI) over an insulin pump (IP1; IP16), or reverse (IP6; IP8), others liked the pump more than the CGM (IP15), with the CGM not working for some (IP3; IP8).

#### Compliance

The degree of independence partly depended on the overall style of self-management between parents and their children, and the compliance with the care regimen. While some young people reported over-management (IP1/IP1P; IP5), others were not following care recommendations strictly (IP2). The omnipresence of the disease and the devices was reported as overwhelming for some participants who were strict in their management (IP5P); especially in puberty, participants reported that it was hard to control blood glucose (IP14; IP15-16P), and that they made use of the devices to improve self-care (IP14).

#### Dignity and empathy/feedback

Negative self-management outcomes impacted participants’ dignity related to their sense of pride and self-respect, for example when unfair treatment because of diabetes occurred. A sense of discrimination was reported by some participants about being unfairly treated at school (IP14P).

*“I was forced to go back in sickbay which I didn’t want to go there because the stomach bug was there and that’s really bad for diabetics to get a stomach bug. So we had to actually go to the hospital and change my claim. that I am allowed to inject in class*.*” (IP14)*

One parent said that *“we had to go through a lot of steps [to use the CGM in class] you sort of feel like there’s this constant discrimination for something that he has no control over… and [there are] safety concerns”* (IP3P). In another situation “*they sort of buddied them up for the first school camp but I think they don’t have to be coupled just because they’ve got type 1 diabetes”* (IP13P); especially young people’s dignity could be impacted. Despite these challenges, empathy was reported – most young people explained that their friends, peers, and family members accepted their medical condition and were very supportive (IP1; IP5P; IP13). Empathy was also expressed by the healthcare team when feedback and support were reported (*see facilitating conditions*). Parents or the healthcare team provided feedback based on data sharing, as well as devices in the form of automated feedback.

#### Hope and joy

Overall, the majority of participants hoped and expected improvement in their management with the devices, and tried to achieve normality in life, being able to enjoy life rather than suffer from the omnipresence of the disease (IP5P; IP15-16P). Diabetes burnout was mentioned as a challenge with the omnipresence of diabetes technologies including constant messages (IP4P) and the burden of wearing the pump all the time (IP5P). Participants described high psychological pressure related to diabetes management, including anxiety (IP1; IP2P; IP4; IP8P; IP14). Use of devices helped to alleviate this anxiety, especially for parents (IP1-2P; IP7; IP12P; IP16P). Moreover, participants tried to manage negative feelings such as discomfort, annoyance, and frustration related to device insertion and site changes (IP2; IP14; IP16), carrying several devices (IP3; IP14P; IP16), and operating the devices (IP3). In particular, pump tubing was mentioned as cumbersome (IP1; IP4; IP7; IP9). Breath devices, such as breath ketone sensors, were considered a potentially interesting non-invasive alternative to reduce pain related to needles and finger pricking (IP1). New CGM and pump models were expected to solve these challenges (IP14), for example with fewer calibration requirements (IP2P), or easier insertion expected for the new CGM models (IP16). Overall, participants perceived that “benefits outweigh the negatives” (IP3P) regarding diabetes technologies.

#### Privacy

Privacy concerns were not reported throughout the interviews, none of the participants mentioned data privacy related aspects.

Overall and to summarize, expectations of what devices should look like were mentioned throughout the interviews and in accordance with all theoretical factors from the models. Thus, expectations could be considered an overarching theme/category. The summary of expectations resulted in a list of ideal device characteristics (mainly related to CGM and insulin pump use) including specific features and designs, presented in **Table 3**. These included, for example, improved reliability and accessibility of diabetes technologies, facilitated device interconnectivity, data sharing and fully automated closed loop systems, improved device algorithms, device non-invasiveness, reduced device sizes and the number of devices to be carried.

**Table 3.**
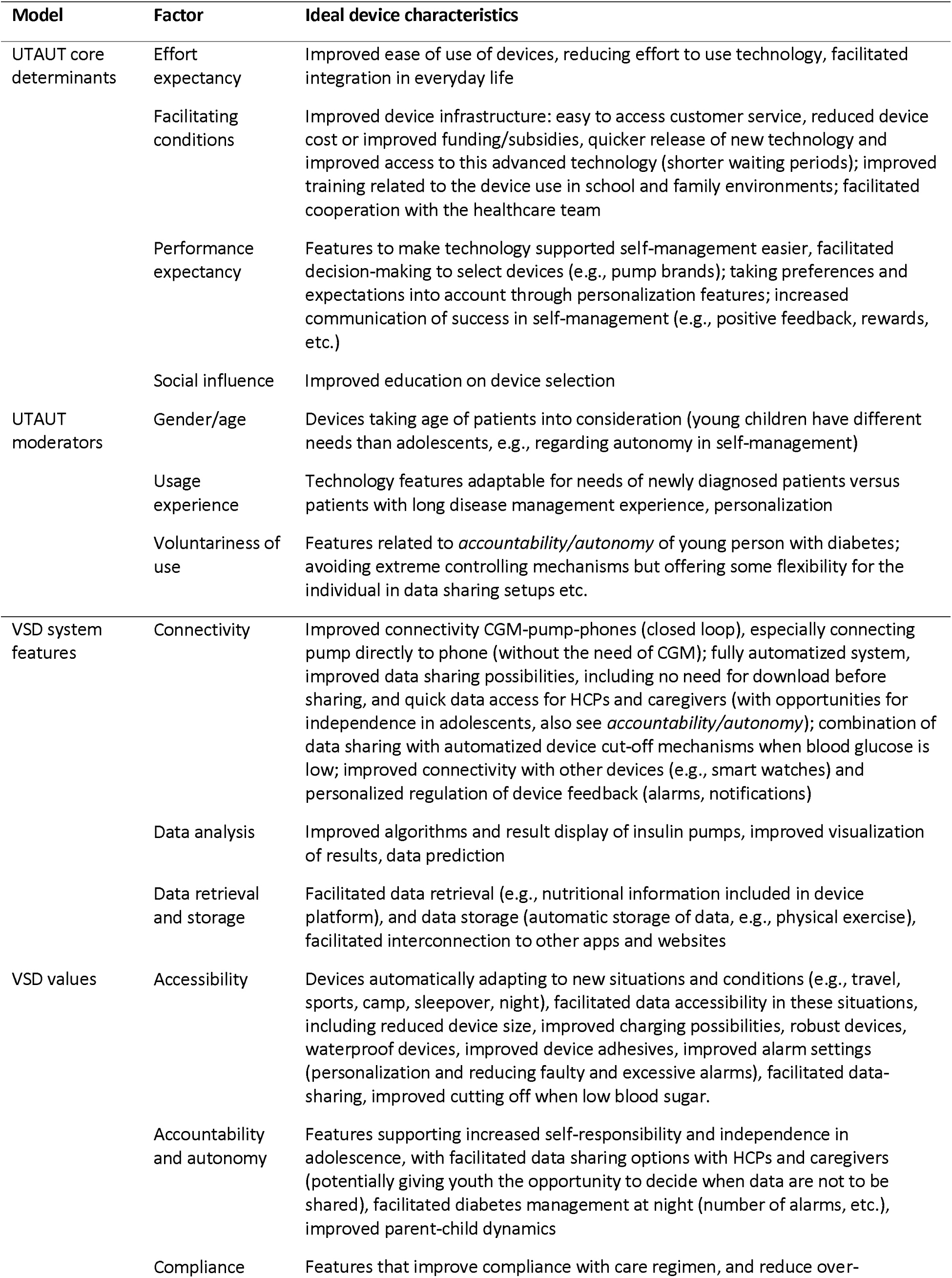
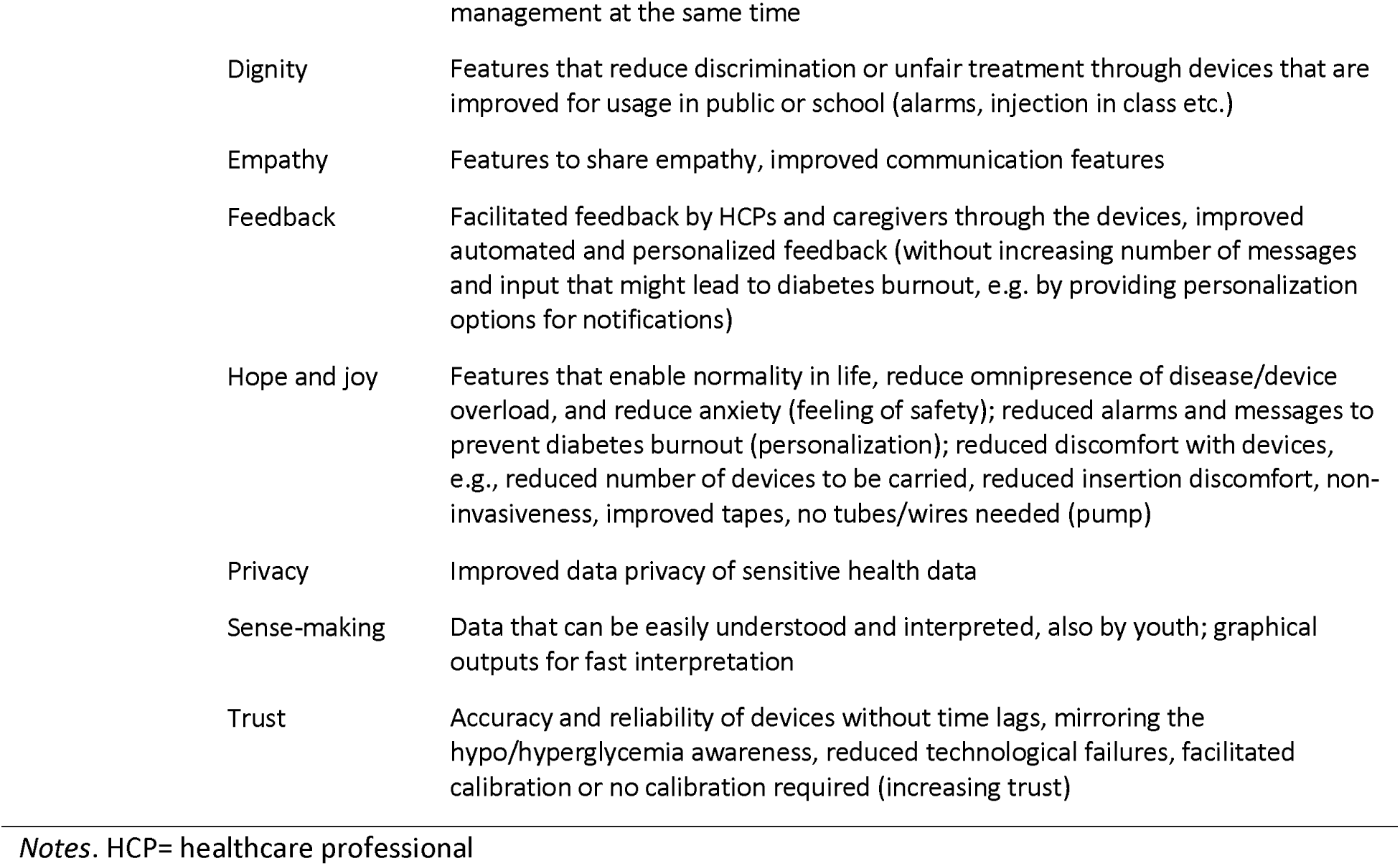
Ideal Device Characteristics based on Theoretical Factors and Interview Findings

## Discussion

All of the factors in our hybrid theoretical framework, except one (privacy), were addressed in the analysis of the interview data, indicating that this framework is a useful foundation for structuring the analysis and findings. This also demonstrates the alignment of the empirical interview data with both theoretical models. We were intrigued by the fact that participants in our study did not raise issues of privacy, as this has been considered of great importance in previous research examining VSD of technologies [27]. Future research should focus specifically on privacy aspects to elucidate potential concerns of young people and their parents, or why this is not an issue of concern for them.

We compared the current study findings with a previous systematic integrative review of 17 studies on young people and their caregivers’ experiences of using technologies to manage T1DM [8]. The review identified eight themes 1. expectations of the technologies prior to use, 2. perceived impact on sleep and overnight experiences, 3. experiences with alarms, 4. impact on independence and relationships, 5. perceived usage impact on blood glucose control, 6. device design and features, 7. financial cost, and 8. user satisfaction. Despite independent analysis of both studies, there was a major overlap between the review themes, and our UTAUT-based (**Figure 1**) and VSD-based (**Figures 2 and 3**) study findings.

**Figure 1.**
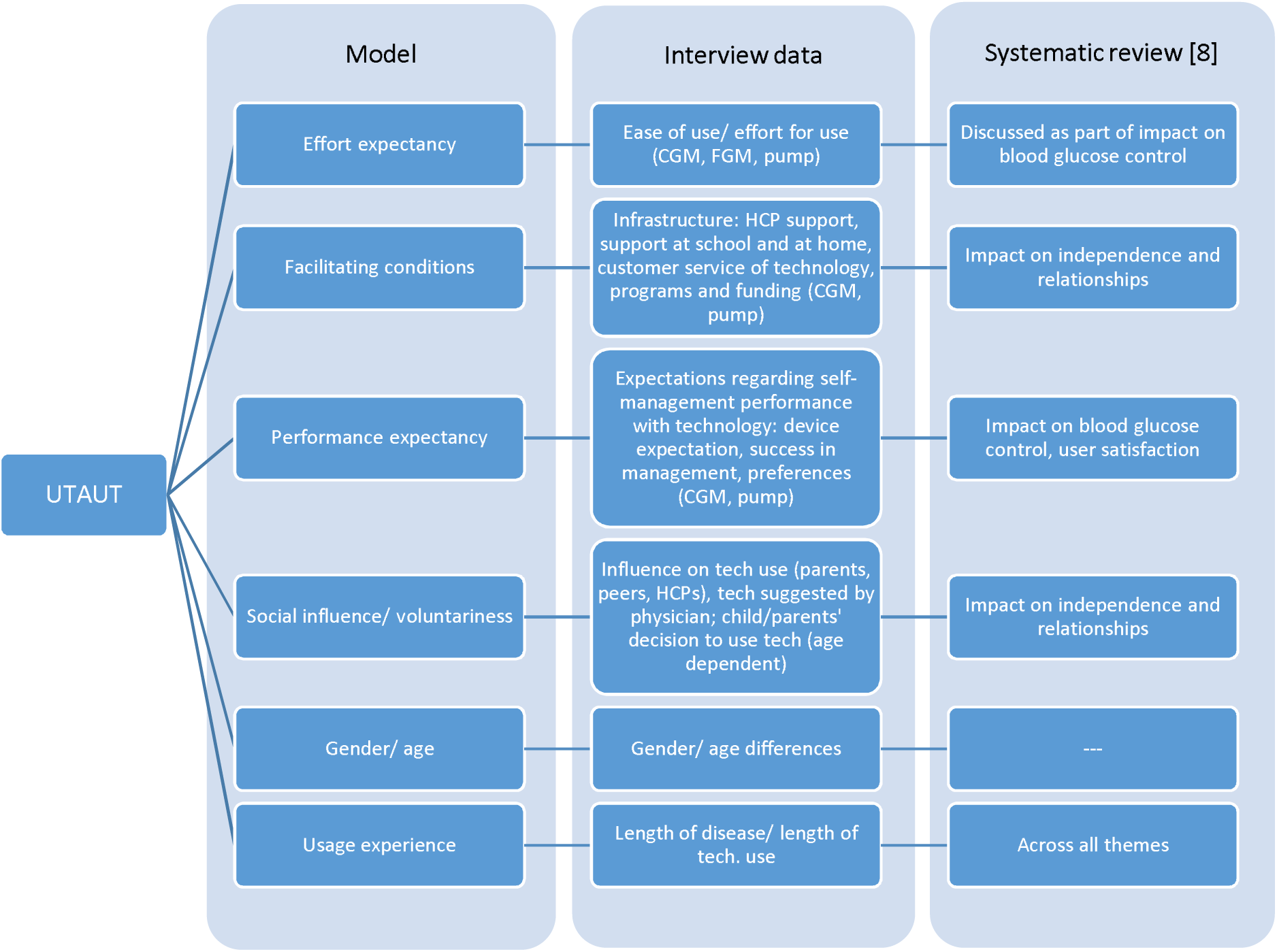
Overlap between UTAUT, interview data and systematic review [8]. *Notes*. CGM=continuous glucose monitoring, FGM=flash glucose monitoring, HCP=healthcare professional

**Figure 2.**
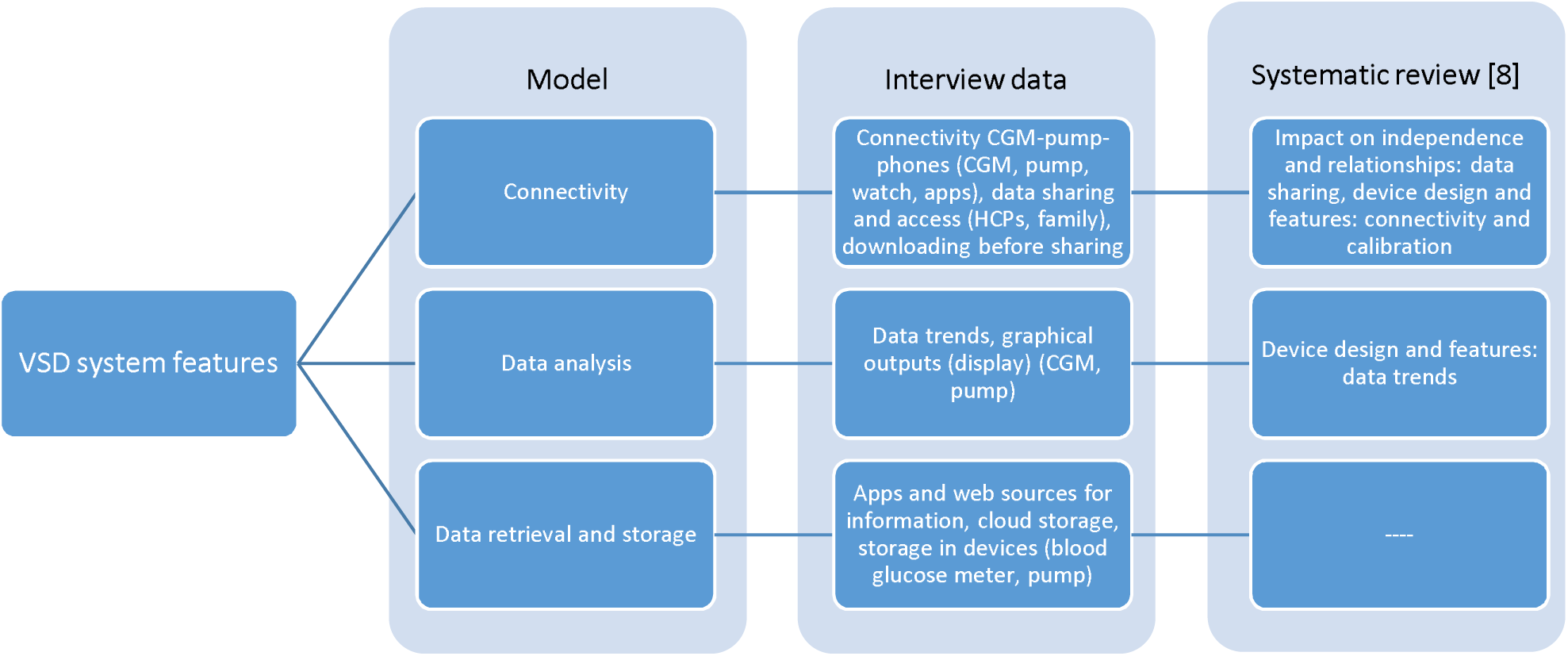
Overlap between VSD system features, interview data and systematic review [8]. *Notes*. CGM=continuous glucose monitoring, HCP=healthcare professional

**Figure 3.**
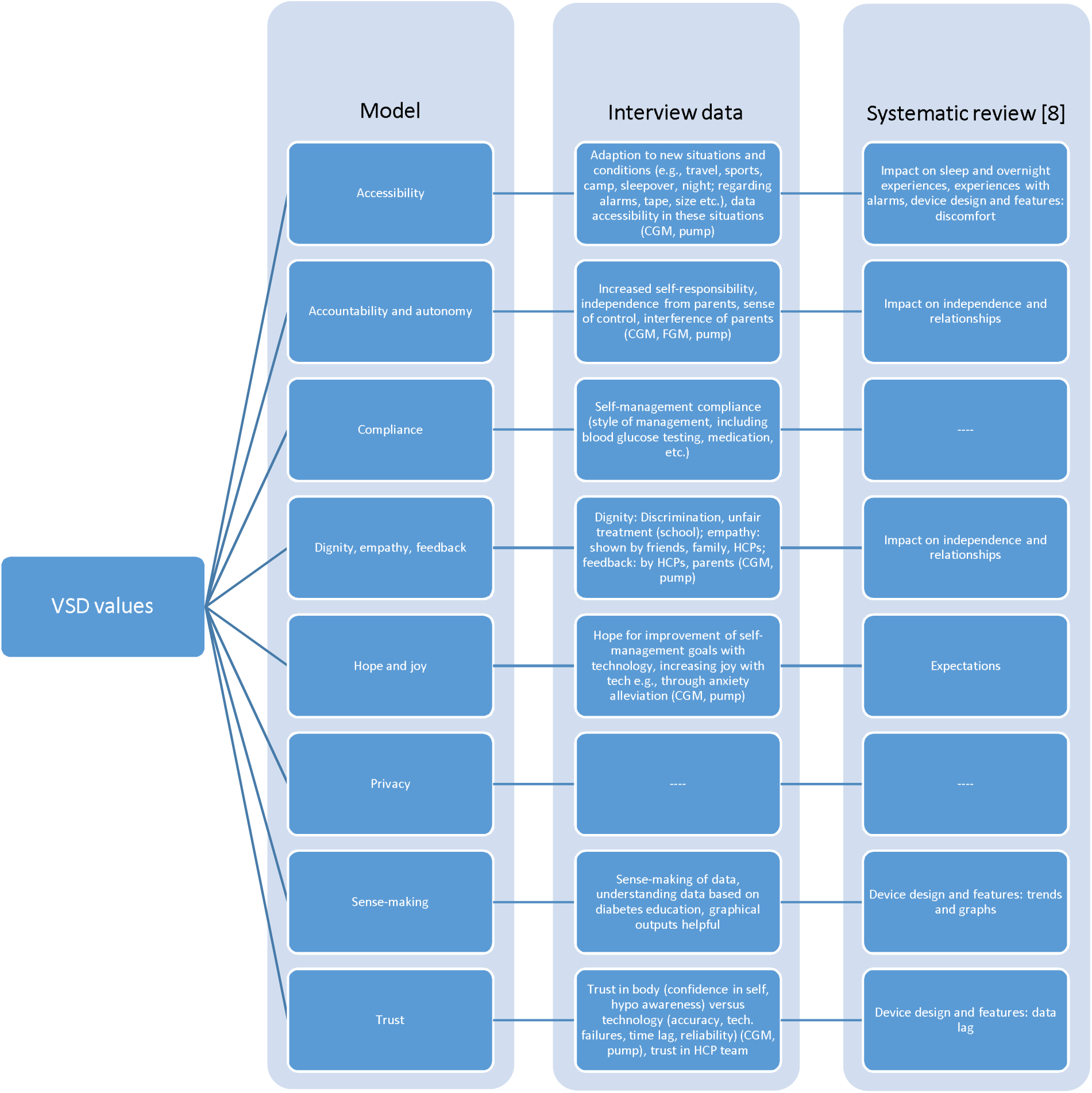
Overlap between VSD values, interview data and systematic review [8]. *Notes*. CGM=continuous glucose monitoring, FGM=flash glucose monitoring, HCP=healthcare professional

Messer [28] argues that with new technological advancements, expectations about new diabetes technologies are high in some individuals at first (idealism), but then fall when reality does not match these expectations. The systematic review [8] reported that some of these expectations related to the self-sufficiency of these technologies, resembling an actual artificial pancreas system which can make life easier and enable normality, also reducing the burden of the disease. Similar wishes and expectations were expressed in the interviews, relating for example to fully automatized systems (*factor connectivity*). In line with the systematic review [8], participants in our study indicated that reality diverted from these expectations, with inaccuracy problems reported for CGMs (time lag in interstitial fluid measurements), and technical failures occurring in both CGMs and insulin pumps. When reality does not match initial expectations this can lead to risk of non-adherence and discontinuation of therapy due to frustration [28]. On the other hand, not all users initially set their expectations high as shown in another study [29], with some people expecting inconveniences regarding technical limitations, cost, wearability, or similar aspects before using the technologies [29]. Overall, accuracy and reliability were highlighted as the most important technology criteria in our study in line with other studies [8, 30].

Apart from expectations prior to use, diabetes management at night and device alarms as found among the review themes [8] were major concerns for participants in our study (factor *accessibility*), while independence was a topic especially raised by adolescents/teenagers (factor *accountability/autonomy*) in both our study and the review. Similarly, Babler and Strickland [31] found that adolescents experienced challenges with independent care and conflicts with their parents. Diabetes-related distress, family conflict, and depressive symptoms are reported barriers towards using diabetes technologies [32]. Previous research describes a learning curve traversed by newly diagnosed individuals as they gradually learn how to self-manage T1DM with devices and in cooperation with important others such as the healthcare team and parents [33]. Distress was mentioned in our study as being particularly high in the early days following diagnosis. Both the review [8] and our study reported that diabetes technologies were able to alleviate psychological challenges such as anxiety to some extent.

Outcomes of technology use for self-management and overall satisfaction with the devices were discussed as part of the UTAUT factors *performance/effort expectancies* and VSD values in our study, with most participants acknowledging the benefits of the devices. A previous study on CGM and insulin pump use in the US and in Germany [34] stated that 47% of pump users were very satisfied with the pump and 98% would recommend the pump to others, whereas only 84% would recommend CGM to others. Apart from device failures, and in line with the review [8], participants in our study reported that cost and funding were major barriers to device accessibility.

Finally, participants highlighted certain aspects beyond the themes of the systematic review [8]. This included perceived discrimination due to having a chronic disease such as T1DM. More research is needed that examines disease-related discrimination. In contrast to a previous study showing difficulties in integrating technologies into clinical workflows [30], most participants in our study reported a smooth process of sharing their diabetes data with the healthcare team and integration of these data into consultation. According to Vrijhoef, de Belvis [35], Integrated Care Pathways (ICPs) can be used for mutual decision-making between patients and healthcare professionals, supported by information technologies which facilitate patient empowerment and improve monitoring and management [35].

Overall, one particular strength of our study included the combination of data-driven and theory-driven analysis. None of the 17 studies included in the systematic review [8] used a theoretical foundation to underpin their examination of experiences despite the proven benefit of the use of theory in research [10]. Incorporating knowledge from two different theoretical paradigms (technology adoption and technology design) into our study of technology experiences and preferences has enabled us to produce research supported by a solid foundation, by combining new results with existing knowledge. It also addresses a general lack of a theoretical basis in studies of diabetes and other health technologies; this may be due to available well-established practice guidelines for managing diabetes that are used instead of theory [9]. Moreover, the combination of the UTAUT with VSD allows us to bring together two theoretical approaches based on different paradigms. In this way, weaknesses of each approach can be compensated for, and a comprehensive foundation is now available for the advancement of diabetes technologies.

## Study Limitations

Our findings were based on self-report of young people with T1DM and their caregivers. The additional perspectives of healthcare professionals would also provide valuable insights into this topic. A degree of self-selection of the participants was unavoidable due to the voluntary nature of study participation. This might have led to an overrepresentation of young people with T1DM who managed their disease well. Perspectives might differ in people with T1DM who struggle in their management, or who do not follow their care regimen.

## Conclusion

Our study findings indicate that technologies for diabetes self-management require continual advancement to meet the needs and expectations of young people with T1DM and their caregivers. Understanding their experiences, and challenges with the devices, enabled us to identify ideal device characteristics that can be useful in designing and developing improved technologies, ideally including participatory design approaches. In our study, theoretical technology adoption and value sensitive design approaches proved useful as a combined foundation for structuring study findings regarding technological experiences and influencing factors. A combination of both approaches is suggested to advance the design of diabetes technologies.

## Data Availability

All data produced in the present study are available upon reasonable request to the authors.

## Acknowledgements/Funding

This research was funded by and has been delivered in partnership *with Our Health in Our Hands*, a strategic initiative of the Australian National University, which aims to transform healthcare by developing new personalized health technologies and solutions in collaboration with patients, clinicians, and health care providers. Moreover, this work was supported by a postdoc fellowship of the *German Academic Exchange Service DAAD* (NBS). We are grateful for the support of Dr Antony Lafferty, Dr Maria Cecilia Garcia Rudaz, and staff and patients at the Canberra Health Services Paediatric Diabetes Service. AT gratefully acknowledges the support of the Australian Research Council for a Future Fellowship (FT200100939) and Discovery grant DP190101864. AT also acknowledges financial support from the North Atlantic Treaty Organization Science for Peace and Security Programme project AMOXES (#G5634).

## Author’s Contributions

NBS, AP, JD, and MC had full access to all the data in the study and take responsibility for the integrity of the data and the accuracy of the data analysis. All authors were involved in study concept and design. HS and AH guided the development of the theoretical foundation of the study. MC and NBS conducted the initial analysis. Findings were discussed with AP. MC, NBS, AP, and JD were involved in the subsequent analysis and interpretation of the data. NBS drafted the manuscript; all authors were involved in revision. JD and HS supervised the study.

## Data availability

Further information about the study can be provided upon request (corresponding author).

## Ethical approval

Ethical approval was obtained by the Australian National University’s Human Ethics Committee and ACT Health Human Research Ethics Committee (HREC, Australia) in October 2019 (2019.ETH.00143; 2019/ETH121700). In addition, a variation for online interviews was approved by the same committees in April, 2020. All interviews were based on informed consent.

## Abbreviations

CGM: Continuous glucose monitoring
FGM: Flash glucose monitoring
IP: Interview participant
T1DM: Type 1 Diabetes Mellitus
TAM: Technology Acceptance Model
TRA: Theory of Reasoned Action
UTAUT: Unified Theory of Acceptance and Use of Technology
VSD: Value Sensitive Design
WAM: Web Acceptance Model

## Footnotes

1 We focused on this age group, as first, government subsidies are limited to those aged under 21 [23]. Second, younger individuals have been reported to be “more exposed to new technologies and easier to absorb the new technological advancements with minimum effort” [24, p. 2]. And third, adolescence – with a transition from childhood to adulthood – is accompanied by general life challenges affecting diabetes management [25, 26].

