## Appendix 1 for "Diabetes Technology Experiences of Young People Living with Type 1 Diabetes Mellitus and their Parents: Hybrid Theoretical Foundation Guided Analysis"

Appendix 1. Initial Themes Derived from the Interviews

| 1 Sociodemography |  |
| --- | --- |
| 2 Diabetes and management | Diabetes education |
|  | Diagnose |
|  | Family members with diabetes |
|  | Feelings about diabetes (anxiety, confidence in self, discrimination) |
|  | Hyperglycemic events |
|  | Hypogylcemia (un)awareness |
|  | Other diseases |
|  | Style of management (Independence, struggles, overmanagement) |
|  | Support (friends, peers, HCPs, parents, school) |
| 3 Device use | Additional devices used (Apple watch, apps, finger pricking devices, ketone sensors, web) |
|  | Device brands used |
|  | Influences on device use (peers, parents, HCPs) |
|  | Length and frequency of device use |
|  | Preferences |
|  | Waiting lists & programs |
| 4.1 Device types: CGM |  |
| 4.1.1 Feelings related to CGM | Negative feelings |
|  | - Annoyance/discomfort/frustration |
|  | - Anxiety (anxiety alleviation, parental stress) |
|  | - Boredom |
|  | - Discrimination |
|  | - Embarrassment |
|  | - Lacking customer service |
|  | Positive feelings |
|  | - Making self-management easier |
|  | - Sense of Control/confidence/role model/freedom |
|  | - Success in management |
|  | - Trust |
|  | Other feelings |
|  | - Device expectation |
|  | - Independence and parental interference |
|  | - Omnipresence |
|  | - Preferences |
| 4.1.2 Tech. characteristics | Accuracy and reliability |
|  | Alarms |
|  | Calibration |
|  | Carry devices (amount of devices, taking devices off, waterproof) |
|  | Changing sensor |
|  | Connectivity |
|  | Cost |
|  | Customization |
|  | Data sharing |
|  | Device screen and trends |
|  | Device size |
|  | Ease of use |
|  | Hypo prevention & awareness |
|  | Insertion site |
|  | Life span |
|  | Needles |
|  | Tapes |
|  | Tech. failures |
|  | Time lag |
| 4.2 Device types: FGM | Feelings related to FGM (sense of control, confidence, role model) |
|  | Tech characteristics (ease of use) |
| 4.3 Device types: Insulin pump |  |
| 4.3.1 Feelings related to pump | Negative feelings |
|  | - Annoyance/cumbersome/burden |
|  | - Anxiety (anxiety alleviation, parental stress) |
|  | - Bribing |
|  | - Difficulty decision making |
|  | - Dislike cost and funding |
|  | - Embarrassment |
|  | - Envy |
|  | Positive feelings |
|  | - Freedom |
|  | - Making management easier/convenience |
|  | - Safety net |
|  | - Sense of Control/confidence/role model |
|  | - Success in management |
|  | - Trust |
|  | Other feelings |
|  | - Device expectations |
|  | - Preferences |
|  | Situations (camp, school, sports) |
| 4.3.2 Tech. characteristics | Accuracy & Reliability |
|  | Alarms & night |
|  | Calculation |
|  | Calibration |
|  | Carry devices (taking device off, travel with pump, waterproof) |
|  | Changing pump |
|  | Closed loop |
|  | Cost & funding |
|  | Customer service |
|  | Data sharing & storage |
|  | Display |
|  | Ease of use |
|  | Insertion site |
|  | Insulin |
|  | Interconnectivity |
|  | Medtronic |
|  | Needles and injections |
|  | Size |
|  | Tape & patches |
|  | Tech failures |
|  | Tubes pump |
