## Appendix 2 for "Diabetes Technology Experiences of Young People Living with Type 1 Diabetes Mellitus and their Parents: Hybrid Theoretical Foundation Guided Analysis"

Appendix 2. Alignment of Initial Themes with Theoretical Factors ­– Data Excerpts

| **Model** | **Factor** | **Interview data themes** | **Data excerpts** |
| --- | --- | --- | --- |
| UTAUT core determinants | Effort expectancy | Ease of use/effort for use (CGM, FGM, pump) | IP1: "a prick... takes a bit of work to get ready and set up"  IP5: "when like I'm in a rush"  IP4: "you just wipe it [FGM] and you get the number"  IP3: "less thinking for yourself"  IP7: "When you just click that, so you can bolus there, put a basal on, like all that cool stuff"  IP5P: "needles, which is a different way of life"; IP9: "easier to do a bolus" (pump) |
|  | Facilitating conditions | Infrastructure: HCP support, support at school and at home, customer service of technology, programs and funding (CGM, pump) | IP14P: "we had to wait a year"  IP13P: "we're waiting four years"  IP8P: "we went on a waiting list months ago, years ago"  IP15P: "the hospital had organised a loan one, and it was a Medtronic"  IP14P/IP5P: delays caused by the Australian Therapeutic Goods Administration, general technology release delays  IP5P: "that kind of process would be very helpful for families, I think, ... just trying to get that health fund and device marrying up"  IP5P: "you sort of get to that four-year window and then you hope for the rest of that remaining year, and you prepare yourself that if that pump dies, that you can revert back to insulin needles if needed"  IP2P: "we rang up in complaints and no one would listen to us" (customer service)  IP13: "You have to ring a number which goes through to India ... and you can be on the phone for quite a long time, and invariably, it's at a time that's not good and, you know, it's stressful"  IP14P: "there is a hotline, ring before four or five o'clock, the hospital"  IP4: "we can call them [healthcare team] like at any time of the day"  IP16P: "easy to access" (hospital)  IP16P: "because they check your heart rate... if there are any lumps or anything... a doctor needs to see that"  IP2P: "we got so much help here... which you don't get in the country"  IP15P: "it was like they [paediatrician] opened up a textbook... instead of treating each kid individually" (rural clinic)  IP15P: "the paediatrician, everybody was separate and nobody communicated. And then we thought we'd try ***... they have meetings... so everyone's on the same page" (rural versus urban clinic)  IP13P: "I don't think that [diabetes] is her specialty so I don't go there" (GP)  IP3: "weren't accepting the CGM" (school)  IP4: "there was one time where a teacher asked to take the pump... well, one of my diabetic friends that goes to that school, he got his pump off him for the day, which probably wasn’t good"  IP3P: "they [hospital team] wrote a letter to the school"  IP6P: "otherwise we're phoning the school and saying 'can you go and get him out of class'"  IP16P: "it [CGM] made it easier for teachers" |
|  | Performance expectancy | Expectations regarding self-management performance with technology: device expectation, success in management, preferences (CGM, pump) | IP14: "weekly summary of how you are tracking"  IP1: "satisfaction of seeing it successful"  IP6P: "he loves it" |
|  | Social influence/ voluntariness of use | Influence on tech use (parents, peers, HCPs), tech suggested by physician;  child/parents' decision to use tech (age dependent) | IP1P: "[physician name] sort of seems to be quite keen on devices and quite keen on pumps"  IP1: "all the kids have Dexcom, my age"  IP14: "they [peers] all have Dexcom"  IP10P: "we made the choice that [name] was to enjoy her childhood, and we would manage her diabetes"  IP3P: "I just tell him the carbohydrate value... and then he makes the decision after that"  IP16P: "I don't want to force her since it's her body" |
| UTAUT moderators | Gender/age | Gender/age differences | ––– |
|  | Usage experience | Length of disease/length of tech. use | IP5P: "we struggled ourselves" (diagnosis)  IP3P: "[diagnosis] was really really emotional and very distressing"  IP4P: "Google it and see what it is all about"  IP6P: "it was hard work, but it's got a lot easier as we've gone on" (learning)  IP3P: "now that they're very in the routine and it's part of our lives, I'm very grateful for my experiences" (struggle at beginning) |
| VSD system features | Connectivity | Connectivity CGM-pump-phones (CGM, pump, watch, apps), data sharing and access (HCPs, family), downloading before sharing | IP14P: the "devices ... talk to each other"  IP7P: "we're using the Dexcom G5, but we're switching... and then there will be, then it's more like a closed loop system"  IP13P: "the icing on the cake" (Apple watch)  IP1P: "get the best of both worlds" (suspend when low and data sharing functions)  IP2P: "they [diabetes systems] all have their pros and cons. A bit like Ford and Holden"  IP16P: "I can't sleep without CGM" (saftey net)  IP1: "all the kids have Dexcom, my age" (data sharing features lead to popularity of Dexcom) |
|  | Data analysis | Data trends, graphical outputs (display) (CGM, pump) | IP14P: "not always a hundred percent accurate" (using CGM data to calculate a HbA1c) |
|  | Data retrieval and storage | Apps and web sources for information, cloud storage, storage in devices (blood glucose meter, pump) | IP13P: "*** diabetes information and ***... Facebook, I'm part of a group called ***"  IP1P: Facebook  IP4P: "Google it"  IP4: "we've got apps if we need to look up"  IP6: "and the pump, ... it stores all the information that you have" |
| VSD values | Accessibility | Adaption to new situations and conditions (e.g., travel, sports, camp, sleepover, night; regarding alarms, tape, size etc.), data accessibility in these situations (CGM, pump) | IP15P: "he seems to be right so long as he's got his sensor"  IP1P: "having it [device] meant [name] could go to school camp"  IP6P: "he's taken it [the pump] out and just gone to finger pricks on camp"  IP9: "the pump had a critical failure at the camp"  IP6: "taking it [CGM] off for swimming"  IP14P: "sensitive skin"; IP1: "itchy"; IP3: "irritated (adhesives)  IP5P: "just a burden" (carry multiple devices)  IP13P: "you need to put it [pump] on silent otherwise your bag... is making all sorts of wonderful noises" (taking the pump off for sports)  IP15P: "it just constantly alarmed for everything"  IP3P: "it would never stop alarming"  IP1: "very embarrassing, I hate it [alarms in school]" |
|  | Accounta-bility and autonomy | Increased self-responsibility, independence from parents, sense of control, interference of parents (CGM, FGM, pump) | IP1: "I kind of take off the responsibility when I go to sleep" (parents responsible at night)  IP2P: "I'm over it [worries]"  IP15P: "he needs to be doing it [management] more himself"  IP16P: "I don't think I'm ready yet to send her for a sleepover"  IP2P: "it [management at camp] is too big a responsibility"  IP3P: "he doesn't really want to have his parents knowing what he's doing all the time"  IP10P: "she's comfortable doing it herself"  IP15P: "might just be a mum thing" (concerns)  IP2: "it can be annoying because she's texting me a lot"  IP15: "I could live without it [data sharing]"  IP16P: "I can keep an eye on her" |
|  | Compliance | Self-management compliance (style of management, including blood glucose testing, medication, etc.) | IP1/IP1P: "tend to over worry, and overly focus"  IP5P: "sometimes it's nice to have a break" |
|  | Dignity, empathy/ feedback | Dignity: Discrimination, unfair treatment (school); empathy: shown by friends, family, HCPs; feedback: by HCPs, parents (CGM, pump) | IP14P: "she was meant to have gone to the sick bay..., and it just meant she was missing out on quite a lot of school work... it also impacted her social life" (discrimination)  IP3P: "we had to go through a lot of steps [to use the CGM in class]... you sort of feel like there's this constant discrimination for something that he has no control over. And safety concerns" (discrimination)  IP13P: "they sort of buddied them up for the first school camp... but I think they don't have to be coupled just because they've got type 1 diabetes" (discrimination)  IP5P: "they [siblings] were all lining up for finger pricks" (empathy)  IP13: "all of them know [class mates]... they're always supportive"; IP1: "my friends know" (empathy) |
|  | Hope and joy | Hope for improvement of self-management goals with technology, increasing joy with tech e.g., through anxiety alleviation (CGM, pump) | IP15P: "sometimes he likes to have a break" (hope for normality, enabling joy)  IP16P: "it's a constant watch" (preventing joy)  IP1P: "it's the condition that provokes anxiety, and the device somewhat alleviates it"  IP3P: "benefits outweigh the negatives" |
|  | Privacy | (not mentioned in the interviews) | ––– |
|  | Sense-making | Sense-making of data, understanding data based on diabetes education, graphical outputs helpful | IP16P: "we were learning so much about diabetes"  IP13: "we're still sort of learning today"  IP14: "the endocrinologist... gives me suggestions, we've already tried it"  IP15P: "they [children] are all different" |
|  | Trust | Trust in body (confidence in self, hypo awareness) versus technology (accuracy, tech. failures, time lag, reliability) (CGM, pump), trust in HCP team | IP14: "sometimes it [CGM] gets very inaccurate"  IP14P: "they're [CGM data] not always a hundred percent accurate"  IP16P: "it [CGM]... wears down"..."later in the month"  IP11: "near the end of its [CGM] life"  IP13P: "[name] is quite hypo-aware"  IP10: "falling sensation in your dreams"; IP3: "light headed, shaky"; IP12: "lazy" (hypo feelings) |

*Notes*. UTAUT factors: Venkatesh [12], VSD factors: Dadgar & Joshi [13]; GP= general practitioner; HCP= healthcare professional; IP= interview participant numbers for young people (e.g., IP1), and for parents (e.g. IP1P)
